# Rising rate of non-receipt of vitamin K prophylaxis for newborns, January 2019 – June 2026

**DOI:** 10.64898/2026.08.31.26361837

**Authors:** Nina B. Masters, Karen G. Farrar, Emma Holler, Johnathan M. Lancaster

## Abstract

**Background:** Vitamin K prophylaxis is universally recommended for newborns to prevent life threatening vitamin K deficiency bleeding. Although not on the immunization schedule, vitamin K prophylaxis is often coadministered with hepatitis B birth dose and erythromycin ophthalmic ointment, and rising hesitancy around vaccines/preventive care may spill over into vitamin K administration.

**Methods:** We conducted a retrospective cohort study using Truveta electronic health record data with linked mother-child dyads. Live births among mothers aged 15-49 from January 1, 2019 through June 30, 2026 were included. Vitamin K administration was defined as documentation on the birth date or following day. Logistic regression assessed sociodemographic predictors of non-receipt, and interrupted time series analysis evaluated changes after January 2026.

**Results:** Among 1,026,375 infants, 995,628 (96.97%) had documented vitamin K administration. Non-receipt increased from an average of 2.1% during 2019 –2022 to 4.3% in 2025 and 6.1% in 2026, reaching 8.10% in June 2026. Older maternal age, non-Hispanic or Latino ethnicity, Medicaid or unknown insurance, and year of delivery were associated with greater odds of non-receipt. After January 2026, there was no immediate step change, but the odds of vitamin K receipt declined an additional 10% per month (OR: 0.90; 95% CI, 0.88 –0.91).

**Conclusions:** Vitamin K non-receipt increased over the study period and accelerated after January 2026. Because vitamin K recommendations were not changed by the January vaccine schedule, this association may reflect broader impacts to confidence in newborn preventive care. Future studies should examine causal mechanisms, parental decision-making, and associated clinical outcomes.

---

Vitamin K is essential for the hepatic activation of several coagulation proteins that are essential for normal blood clotting.^1^ Newborns have limited vitamin K stores and are susceptible to vitamin K deficiency bleeding (VKDB), a rare but potentially life-threatening hemorrhagic disorder.^1^ This vulnerability reflects several physiologic factors unique to newborns, including limited transplacental transfer of vitamin K, low hepatic reserves, low concentrations of vitamin K in human breastmilk, and an immature intestinal microbiome with limited capacity for endogenous vitamin K production.^1–3^

In the absence of vitamin K prophylaxis at birth, VKDB may occur from the neonatal period through 6 months of age. Late VKDB, occurring from 1 week to 6 months of life, can be the most serious, presenting with occult intracranial hemorrhage.^1,3^. The Centers for Disease Control and Prevention (CDC) reports that compared to infants who receive vitamin K at birth, those who do not have an 81-fold higher risk of late VKDB.^1^

The American Academy of Pediatrics (AAP) recommends all newborns weighing >1,500 g receive 1 mg of vitamin K by intramuscular injection within 6 hours of birth. Preterm infants with lower birth weights should receive a weight-based dose.^3^ The AAP also recommends that clinicians counsel caregivers on the benefits of vitamin K prophylaxis and the risks associated with non-administration.^3–5^

Despite longstanding clinical recommendations, nonreceipt of vitamin K prophylaxis appears to be increasing in the U.S.^6^ Amid a climate of increased politicization of routine prophylactic measures for infants and general increasing distrust of vaccination and related preventive measures, it is essential to assess whether the non-administration of Vitamin K prophylaxis is accelerating.

Over the past nine months. U.S. health care policy entities have made major changes to longstanding recommendations surrounding routine childhood care, particularly for vaccines. In December 2025, the Advisory Committee on Immunization Practices (ACIP) voted to remove the universal birth dose recommendation for Hepatitis B vaccination, and in January 2026, the Department of Health and Human Services released a revised childhood immunization schedule, without holding a corresponding ACIP vote, to r educ e recommended routine vaccines from 17 diseases to 11. ^7–9^ Federal vaccine recommendations became more complicated o n March 16, 2026, when a federal district court stayed the revised schedule and all votes undertaken by the appointed members of the sitting ACIP.^10^

Although the recommendations surrounding vitamin K prophylaxis did not change, vitamin K refusal has been shown to be strongly associated with refusal of other newborn preventive measures for which recommendations did change, including Hepatitis B vaccination.^11–13^ The changes to the vaccine schedule over the past year, along with pending questions of their legality, has generated substantial media attention, which has most likely led to increased public awareness of these routine public health prevention strategies.

In the aftermath of changes to the vaccine schedule, the AAP and CDC diverged in their recommendations for childhood vaccinations, which likely created additional confusion for parents. We sought to assess whether changes to the vaccine schedule may have impacted preventive health measures for infants beyond the vaccine schedule itself – specifically intramuscular vitamin K prophylaxis at birth.

## Methods

This cohort study used a subset of Truveta Data, an electronic health record (EHR) database with over 130 million individuals from a collective of healthcare systems across the US. ^14,15,16^ Truveta Data include structured demographics, details of healthcare encounters, diagnoses, medication administrations, procedures, charge item data, and a deterministic linkage between a subset of mothers and children. Data undergo syntactic and semantic normalization, are de-identified by expert determination under the HIPAA Privacy Rule (45 CFR §164.514[b]), and thus are exempt from Institutional Review Board review due to classification as non-human participant research.

For this study, we examined data for mothers with evidence of live birth and delivery dates between January 1, 2019, and June 30, 2026, who were aged 15 to 49 years at the time of delivery. For individual mothers who had given birth multiple times during the study period, each live birth was counted separately as a unique event if at least 300 days had transpired between births.

We classified vitamin K administration in the infant EHR (codeset provided in Supplementary Table S1) if vitamin K was documented on the child’s birthdate or one day after to allow for differences in documentation practices. Maternal insurance type was determined by selecting the insurance type billed closest to (and within 30 days of) of the child’s birth. We examined time trends of vitamin K non-receipt over the study period, performed multivariable logistic regression to assess how maternal socio demographic factors and newborn sex were associated with non-receipt of vitamin K, and performed an interrupted time series analysis to evaluate the impact of the January 2026 change to the pediatric immunization schedule. Models used cluster-robust standard errors to account for clustering by mother, as some mothers had multiple children over the study period. Analyses were performed in R, version 4.4.1, within a cloud-based notebook environment leveraging Apache Spark, version 3.5. P-values less than 0.05 were considered statistically significant.

## Results

We identified 1,026,375 liveborn infants born to 825,599 unique mothers with birth dates between January 2019 and June 2026 that met inclusion criteria. Among these, 995,628 newborns had documented vitamin K administration within one day of birth, corresponding to 96.97% of live births across the full study period.

### Rate of non-receipt of vitamin K prophylaxis

During the study period, the rate of non-receipt of vitamin K increased over time. While rates averaged 2.1% from 2019-2022 and increased modestly (0.1-0.3 percentage points per year) over this four-year period, non-receipt rates began to increase sharply in 2025, reaching 4.3%, and climbing to 6.1% in 2026 (Table 1, Supplementary Figure S1). Looking at monthly trajectories of vitamin K non-receipt, 5.15% of births did not have documented receipt of vitamin K in January 2026, increasing to 8.10% in June 2026 (Figure 1A). Rates of vitamin K non-receipt were similar for newborns of male and female sex from 2019 to 2023 (averaging 0.2 to 0.3 percentage points higher for female newborns) but began to diverge more in 2024 (3.6% for females, 2.7% for males), reaching 6.8% for female and 5.5% for male infants for 2026 (Figure 1B, Supplementary Figure S2).

**Table 1.** Population characteristics of newborns receiving vitamin K prophylaxis at birth, January 2019 – June 2026.

| Characteristic | Vitamin K prophylaxis at birth |  |
| --- | --- | --- |
|  | Received<br>N = 995,628 | Not Received<br>N = 30,747 |
| **Birth Year** |  |  |
| 2019 | 90,398 (98.0%) | 1,826 (2.0%) |
| 2020 | 114,096 (98.0%) | 2,308 (2.0%) |
| 2021 | 137,949 (97.9%) | 2,963 (2.1%) |
| 2022 | 145,671 (97.6%) | 3,645 (2.4%) |
| 2023 | 147,322 (97.1%) | 4,394 (2.9%) |
| 2024 | 149,181 (96.9%) | 4,812 (3.1%) |
| 2025 | 143,046 (95.7%) | 6,379 (4.3%) |
| 2026 | 67,965 (93.9%) | 4,420 (6.1%) |
| **Newborn Sex** |  |  |
| Female | 483,820 (96.7%) | 16,428 (3.3%) |
| Male | 511,808 (97.3%) | 14,319 (2.7%) |
| **Maternal Age** |  |  |
| 15-18 | 10,721 (98.3%) | 181 (1.7%) |
| 19-29 | 397,113 (97.2%) | 11,445 (2.8%) |
| 30-39 | 535,688 (96.9%) | 17,008 (3.1%) |
| 40-49 | 52,106 (96.1%) | 2,113 (3.9%) |
| **Marital Status** |  |  |
| Divorced | 1,862 (97.1%) | 55 (2.9%) |
| Married/Domestic Partner | 481,961 (97.0%) | 15,033 (3.0%) |
| Other | 109 (94.8%) | 6 (5.2%) |
| Unmarried | 242,189 (96.6%) | 8,608 (3.4%) |
| Unknown | 269,507 (97.5%) | 7,045 (2.5%) |
| **Maternal Race** |  |  |
| American Indian or Alaska Native | 2,845 (97.0%) | 88 (3.0%) |
| Asian | 87,947 (98.6%) | 1,292 (1.4%) |
| Black or African American | 116,493 (97.2%) | 3,309 (2.8%) |
| Native Hawaiian or Other Pacific Islander | 10,728 (98.9%) | 121 (1.1%) |
| Other Race | 81,185 (97.1%) | 2,423 (2.9%) |
| White | 69,379 (96.1%) | 2,822 (3.9%) |
| Unknown | 627,051 (96.8%) | 20,692 (3.2%) |
| **Maternal Ethnicity** |  |  |
| Hispanic or Latino | 220,234 (97.5%) | 5,744 (2.5%) |
| Not Hispanic or Latino | 739,410 (96.9%) | 23,375 (3.1%) |
| Unknown | 35,984 (95.7%) | 1,628 (4.3%) |
| **Maternal Insurance** |  |  |
| Commercial | 364,512 (97.1%) | 11,066 (2.9%) |
| Medicaid | 373,850 (96.9%) | 11,864 (3.1%) |
| Other | 60,289 (97.7%) | 1,446 (2.3%) |
| Unknown | 196,977 (96.9%) | 6,371 (3.1%) |

**Figure 1.**
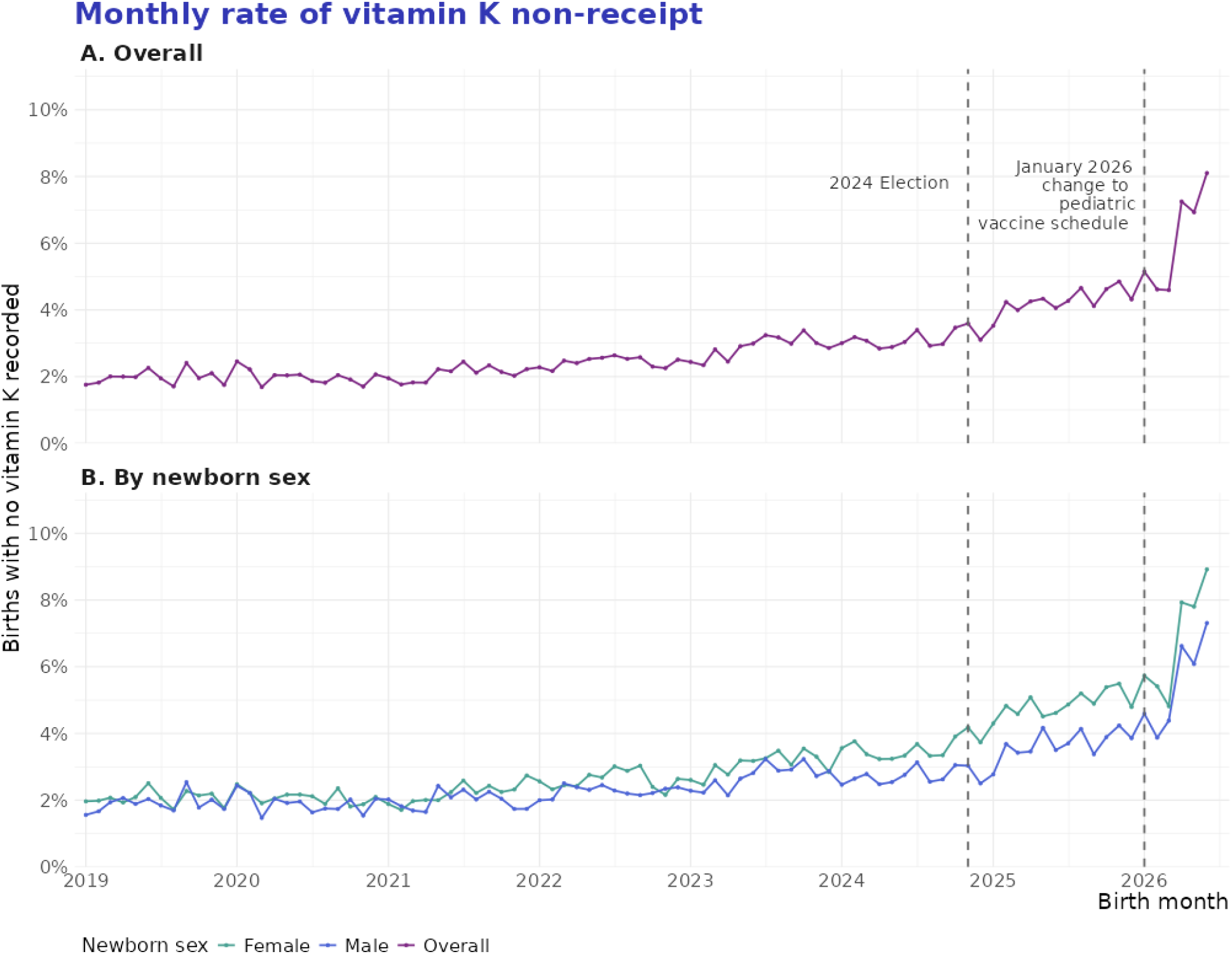
Trends in vitamin K non-receipt over time from 2019 – June 2026 (A) overall, (B) by newborn sex

### Predictors of non-receipt of vitamin K prophylaxis

In adjusted logistic regression analyses, maternal demographics and year of delivery were associated with non-receipt of vitamin K (Figure 2, Supplementary Table S2). Infants born to older mothers had greater odds of non-receipt of vitamin K, with a stepwise association by age (aOR 2.40, 95% CI: 2.05-2.81 for those aged 40-49, aOR 1.86, 95% CI: 1.60-2.16 for those aged 30-39, aOR 1.66, 95% CI: 1.43-1.93 for those aged 19-29; compared to mothers aged 15-18). Infant s born to mothers of non-Hispanic or Latino ethnicity (aOR 1.53, 95% CI: 1.48-1.59) had greater odds of not receiving vitamin K compared to those born to mothers of Hispanic or Latino ethnicity, while infants born to mothers of Asian (aOR 0.40 95% CI: 0.38-0.42) and Native Hawaiian and Pacific Islander (aOR 0.32, 95% CI: 0.26-0.39) race had lower odds of not receiving vitamin K at birth compared to those born to White mothers. Compared to infants born to mothers with commercial insurance, infants born to mothers with unknown (not recorded) insurance had 1.13 times the odds of non-receipt of vitamin K (95% CI: 1.09-1.17) and those with Medicaid had 1.19-fold odds of non-receipt of vitamin K (95% CI: 1.16-1.23). The odds of non-receipt of vitamin K prophylaxis increased by year of delivery, with infants born in 2026 having over 3 times the odds of non-receipt (aOR 3.16, 95% CI: 2.99-3.35) as those born in 2019.

**Figure 2.**
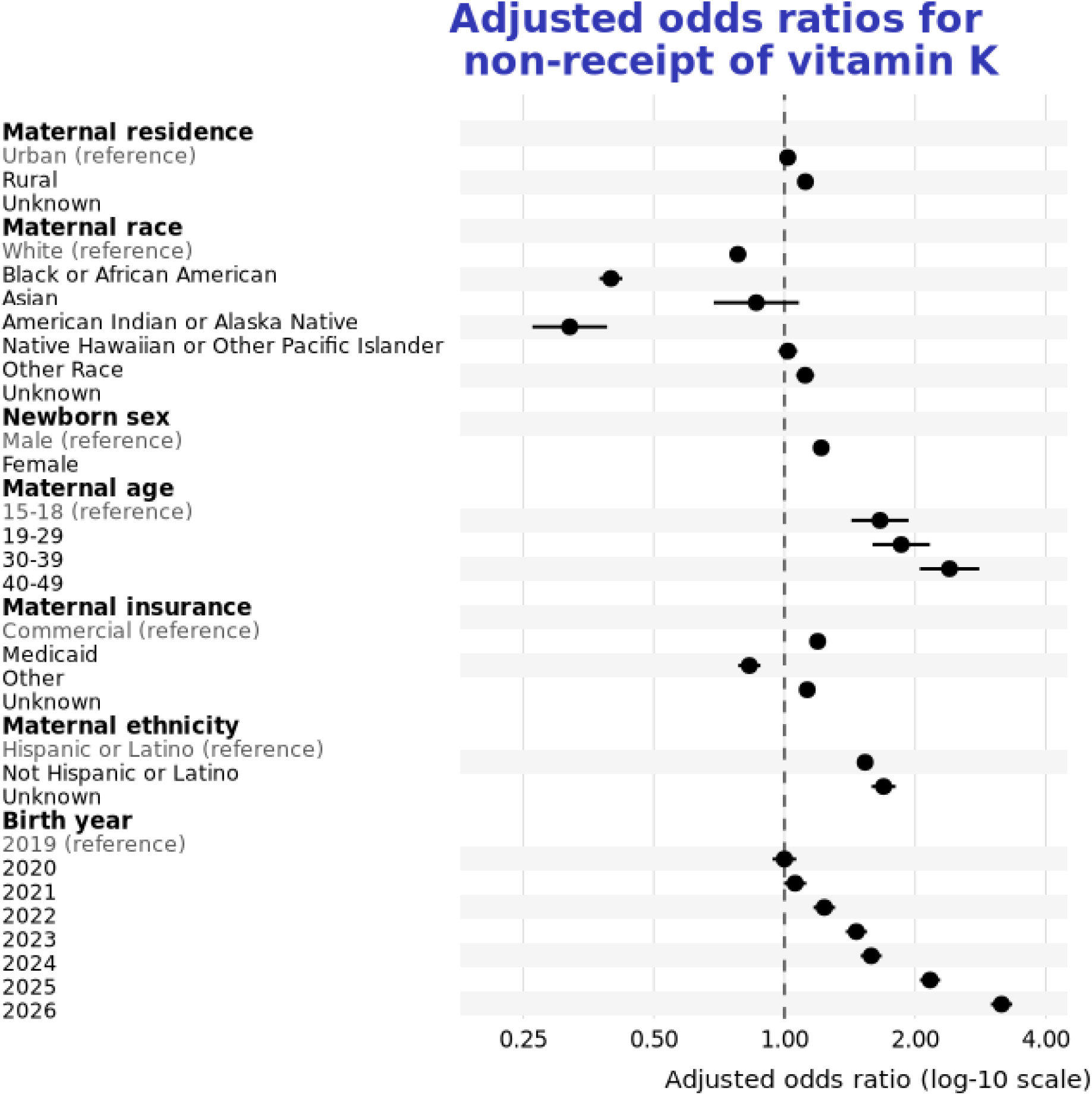
Forest plot of odds ratios for maternal predictors of newborn vitamin K non-receipt

**Figure 3.**
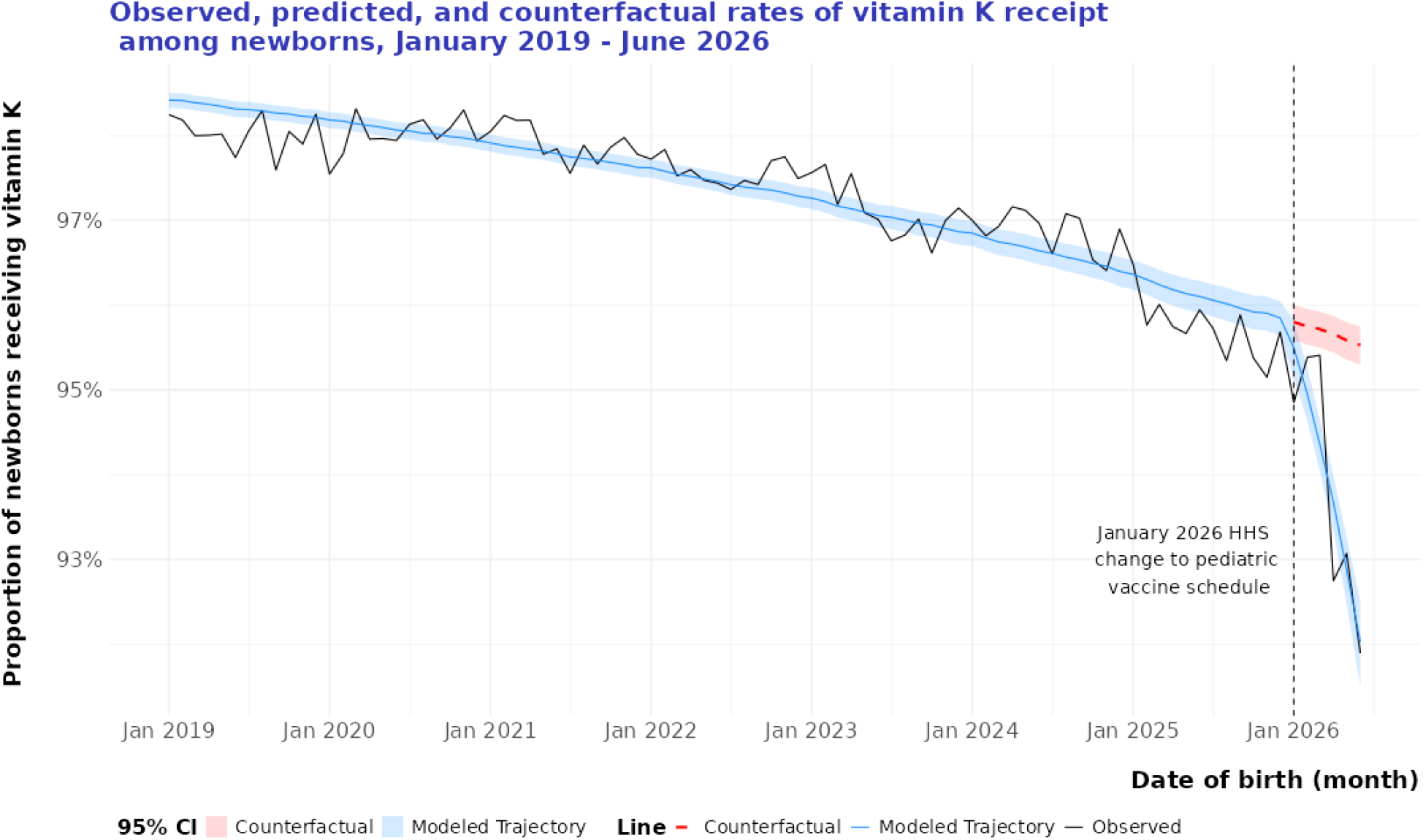
Modeled trajectories of vitamin K receipt using interrupted time series analysis. Figure presents observed data in black, fitted model in blue (with 95% CI) and counterfactual trajectory shown in red in the absence of January 2026 change to pediatric immunization schedule.

### Interrupted time series analysis

From January 2019 to December 2025, receipt of vitamin K prophylaxis declined by an average of 1.0% per month (aOR 0.99, 95% CI: 0.99-0.99). There was no step effect observed immediately after the January 2026 vaccine schedule change (aOR: 1.04, 95% CI: 0.96-1.12), however there was a significant acceleration of declining rates of vitamin K administration – with an additional 10.0% monthly reduction in the odds of receiving vitamin K in the period from January – June 2026 (aOR: 0.90, 95% CI: 0.88-0.91).

## Discussion

The AAP has recommended vitamin K prophylaxis since 1961, and its current guidance continues to recommend intramuscular vitamin K for all newborns within six hours of birth.^3,17^ These 65 years of medical precedent have served to prevent potentially fatal vitamin K deficiency bleeding (VKDB), with global data showing a vitamin K prophylaxis at birth associated with a 98% reduction in late VKDB.^18^ In this large real-world cohort of live births in a healthcare setting from 2019 through June 2026, we found that non-receipt of vitamin K in the immediate newborn period increased over time, with acceleration beginning in 2026. This increase may reflect broader population-level hesitancy for preventive interventions in the post-pandemic period, as well as spillover from a general climate of increased discussion about vaccines, science, and the prioritization of choice in medical interventions.

Our findings support recent published evidence that show rates of non-receipt of vitamin K prophylaxis increasing from ~3% in 2018 to over 5% in 2024 in the U.S.^6^ In our study, we observed several predictors of non-receipt of vitamin K, including increasing maternal age, White race, non-Hispanic or Latino ethnicity and unknown health insurance at the time of delivery. Existing literature supports the finding that increased maternal age ^6,12^, White race, ^6,12^ and female newborn sex ^6,11,12^ are associated with non-receipt of infant vitamin K prophylaxis. These predictors may reflect several overlapping upstream causes, including care setting, local hospital practices, access to prenatal education, and parental beliefs. The disparity seen by newborn sex may reflect an association with circumcision for male newborns, as many hospitals will not perform the procedure in the absence of vitamin K prophylaxis due to increased bleeding risk. ^11^

Looking beyond domestic trends, the phenomenon of increasing vitamin K non-receipt may be a global phenomenon, as a recent study in Sweden found non-receipt had doubled from 2003 to 2021,^19^ and recent reporting in Australia showed rates declined in the two states that conduct vitamin K surveillance, dropping from 98.3% in 2016 to 95.5% in 2025 in Queensland. ^20^

Previous studies have reported potential reasons for non-administration of vitamin K, including a fear of injection, mistrust of medical procedures, or beliefs in alternative ways to prevent vitamin K deficiency bleeding, such as utilization of oral vitamin K. ^12,20^ Research on parental decision-making has found that potential concerns about ingredients, dose, side effects, and information found online can contribute to refusal. This suggests that vitamin K refusal may be part of broader hesitation about newborn preventive care linked to vaccine hesitancy, even though vitamin K is not a vaccine. ^3,13,21^

We hypothesized that changing federal vaccine recommendations may increase confusion and lead to greater mistrust of preventive interventions – even vitamin K, as it is an injection often coadministered with birth dose immunizations and other preventive measures. Our results quantify that since the January 2026 vaccine schedule change, the odds of newborns receiving vitamin K at birth is declining by an additional 10% per month compared to before this schedule change. The January vaccine schedule change *did not* alter recommendations for vitamin K, so any observed association may reflect impacts to parental confidence in routine newborn preventive care and should not be interpreted as a direct causal effect.

While most infants in our study received vitamin K prophylaxis, even small increases in non-receipt can carry significant public health impacts. In 2013, a cluster of VKDB cases in Tennessee occurred among infants whose parents had declined vitamin K prophylaxis. ^22^ Investigators found that some parents declined the injection because of misinformation. ^22^ Four infants developed VKDB; three had intracranial hemorrhage and one had gastrointestinal bleeding. ^22^ If vitamin K refusal continues to rise nationally, more infants will be vulnerable to a preventable and potentially fatal bleeding disorder.

### Strengths and Limitations

This study has several limitations. First, vitamin K administrations were recorded if documented within 1 day of the newborn’s birth date, which does not capture the rate of administration within the 6-hour AAP recommended window. Second, this analysis describes documented vitamin K administration, not parental refusal. Lack of documented administration could reflect refusal, documentation gaps, a newborn being transferred to a different medical facility, data availability challenges, or other clinical circumstances. Third, this study only includes data on births in a medical setting, and thus excludes home births or births in birthing centers. As a result, actual rates of newborn vitamin K non-receipt are likely higher than those reported, as newborns born outside healthcare facilities have much lower rates of vitamin K prophylaxis. ^20^ Despite these limitations, this analysis provides timely evidence on newborn vitamin K administration patterns across a large real-world dataset in the United States. This study helps contextualize a concerning rise in non-receipt of a life-saving prophylactic injection for newborns.

## Conclusions

Vitamin K prophylaxis remains universally recommended by both the CDC and AAP, and its omission places a subset of newborns at risk of life-threatening VKDB. In June 2026, 8% of newborns did not have evidence of vitamin K prophylaxis at birth. If this trend continues, and nonreceipt of vitamin K prophylaxis continues to increase, more cases of VKDB are likely to occur in young infants, likely leading to avoidable morbidity and mortality.

## Data Availability

The data used in this study are available to Truveta subscribers and may be accessed at studio.truveta.com.

## Abbreviations

ACIP: Advisory Committee on Immunization Practices
AAP: American Academy of Pediatrics
CDC: Centers for Disease Control and Prevention
EHR: Electronic Health Record
VKDB: Vitamin K deficiency bleeding

## Contributors Statement

N. Masters conceptualized and designed the study, carried out initial analyses, drafted the initial manuscript, and critically reviewed and revised the manuscript.

K. G. Farrar reviewed analyses and critically reviewed and revised the manuscript.

E. Holler reviewed analyses and critically reviewed and revised the manuscript

J. Lancaster critically reviewed and revised the manuscript.

All authors approved the final manuscript as submitted and agree to be accountable for all aspects of work.

## Acknowledgements

The authors would like to acknowledge Zachary Wallen, PhD, MS and Madhura Vachon, PhD for methodological review and feedback.

## References

1. Frequently Asked Questions About Vitamin K Deficiency Bleeding | Vitamin K Deficiency Bleeding | CDC [Internet]. [cited 2026 Jul 14];Available from: https://www.cdc.gov/vitamin-k-deficiency/faq/index.html

2. Sirachainan N, Komvilaisak P, van Ommen CH, Revel-Vilk S. International Perspectives on Vitamin K Deficiency Bleeding in Infants: A Cross-Sectional Questionnaire-Based Survey. Pediatr Blood Cancer 2025;72(10):e31889.

3. Hand I, Noble L, Abrams SA, Committee on Fetus and Newborn, Section on Breastfeeding, Committee on Nutrition. Vitamin K and the Newborn Infant. Pediatrics 2022;149(3):e2021056036.

4. Ardell S, Offringa M, Ovelman C, Soll R. Prophylactic vitamin K for the prevention of vitamin K deficiency bleeding in preterm neonates. Cochrane Database Syst Rev 2018;2018(2):CD008342.

5. Puckett RM, Offringa M. Prophylactic vitamin K for vitamin K deficiency bleeding in neonates. Cochrane Database Syst Rev [Internet] 2000 [cited 2026 Jul 14];(4). Available from: https://www.cochranelibrary.com/cdsr/doi/10.1002/14651858.CD002776/full

6. Scott K, Miller E, Culhane JF, et al. Trends in Vitamin K Administration Among Infants. JAMA 2026;335(3):272 –4.

7. Kates J, Michaud J. The New Federal Vaccine Schedule for Children: What Changed and What Are the Implications? [Internet]. KFF. 2026 [cited 2026 Jul 14];Available from: https://www.kff.org/other-health/the-new-federal-vaccine-schedule-what-changed/

8. Centers for Disease Control and Prevention. ACIP Recommends Individual-Based Decision - Making for Hepatitis B Vaccine for Infants Born to Women Who Test Negative for the Virus [Internet]. 2025 [cited 2026 Mar 13];Available from: https://www.cdc.gov/media/releases/2025/2025-acip-recommends-individual-based-decision-making-for-hepatitis-b-vaccine-for-infants-born-to-women.html

9. U.S. Department of Health and Human Services. Decision Memo: Adopting Revised Childhood and Adolescent Immunization Schedule [Internet]. 2026 [cited 2026 Jul 14]. Available from: https://www.hhs.gov/sites/default/files/decision-memo-adopting-revised-childhood-adolescent-immunization-schedule.pdf

10. Jenco M. Government appeals AAP vaccine lawsuit ruling; loses bid to delay document release [Internet]. AAP News. 2026 [cited 2026 Jul 17];Available from: https://publications.aap.org/aapnews/news/34981/Government-appeals-AAP-vaccine-lawsuit-ruling

11. Coggins SA, Flannery DD, Mukhopadhyay S, Puopolo KM. Parental Decline of Newborn Vitamin K and Hepatitis B Vaccine Administration by Newborn Sex. JAMA Netw Open 2026;9(6):e2618410.

12. Loyal J, Taylor JA, Phillipi CA, et al. Factors Associated With Refusal of Intramuscular Vitamin K in Normal Newborns. Pediatrics 2018;142(2):e20173743.

13. Hamrick HJ, Gable EK, Freeman EH, et al. Reasons for Refusal of Newborn Vitamin K Prophylaxis: Implications for Management and Education. Hosp Pediatr 2016;6(1):15 –21.

14. Truveta. Our Approach to Data Quality [Internet]. 2025. Available from: https://www.truveta.com/resources/whitepaper/truvetas-approach-to-data-quality/

15. Burkhardt HA, Blach SJ, Burugapalli SR, et al. From fragmented records to living evidence: health system-governed, artificial intelligence-driven, continuously updated real-world clinical data from Truveta. JAMIA Open 2026;9(4):ooag142.

16. Rodriguez PJ, Goodwin Cartwright BM, Gratzl S, et al. Semaglutide vs Tirzepatide for Weight Loss in Adults With Overweight or Obesity. JAMA Intern Med 2024;184(9):1056 –64.

17. Committee on Nutrition. Report of the Committee on Nutrition: Vitamin K Compounds and the Water-Soluble Analogues. Pediatrics 1961;28(3):501 –7.

18. Sankar MJ, Chandrasekaran A, Kumar P, Thukral A, Agarwal R, Paul VK. Vitamin K prophylaxis for prevention of vitamin K deficiency bleeding: a systematic review. J Perinatol 2016;36(1):S29–35.

19. Simatou E, Tsamantioti E, Hallström A, et al. Vitamin K Prophylaxis in Newborns and Bleeding in Infancy. JAMA Pediatr [Internet] 2026 [cited 2026 Aug 12];Available from: 10.1001/jamapediatrics.2026.2606

20. More parents refusing lifesaving vitamin K injection for newborns [Internet]. ABC News. 2026 [cited 2026 Aug 12];Available from: https://www.abc.net.au/news/2026-07-26/parents-refusing-lifesaving-vitamin-k-injection-for-newborns/106938860

21. Rogers TP, Fathi O, Sanchez PJ. Neonatologists and vitamin K hesitancy. J Perinatol 2023;43:1067 – 71.

22. Warren M, Miller A, Traylor J, et al. Notes from the Field: Late Vitamin K Deficiency Bleeding in Infants Whose Parents Declined Vitamin K Prophylaxis — Tennessee, 2013. Morb Mortal Wkly Rep 2013;62(45):901 –2.

23. Marcewicz LH, Clayton J, Maenner M, et al. Parental Refusal of Vitamin K and Neonatal Preventive Services: A Need for Surveillance. Matern Child Health J 2017;21(5):1079–84.

