## supplement for "Rising rate of non-receipt of vitamin K prophylaxis for newborns, January 2019 – June 2026"

### Supplementary Tables and Figures

**Supplementary Table S1.** Vitamin K prophylaxis codes, drawn from medication administration, procedure, and charge item data

| Coding System | Codes | Logic |
|---|---|---|
| RxNorm | 1670191, 1670192, 727624, 727625, 373449, 1161771, 1670374, 317466, 1360307, 412345 | Self and descendants |
| NDC | 69097000396, 00004190706, 52584014000, 51662153702, 69097000367, 46708082605, 62332082610, 00409915750, 46708082610, 680830606, 55154238805, 516621537, 70121168207, 00074915701, 5154395205, 500904521, 62332082605, 516621536, 00004190806, 69097054049, 70121168201, 46066091501, 00548114000, 467080826, 690970709, 11695401401, 690970003, 54868443400, 59346600301, 51662153703, 00409915731, 76329124001, 69097070930, 00409915831, 00409915801, 69097070996, 00409915701, 00409915725, 00409915850, 00409915811, 00409915825, 00409915855 | Self and descendants |
| HCPCS | J3430 | Self and descendants |

**Supplementary Table S2.** Logistic regression results for non-receipt of newborn vitamin K prophylaxis

| Maternal or Newborn Characteristic | Adjusted Odds Ratio | 95% CI |
|---|---|---|
| Birth year | | |
| 2019 | REF | |
| 2020 | 1.00 | 0.94-1.06 |
| 2021 | 1.06 | 1.00-1.12 |
| 2022 | 1.24 | 1.17-1.31 |
| 2023 | 1.46 | 1.39-1.55 |
| 2024 | 1.58 | 1.50-1.68 |
| 2025 | 2.17 | 2.05-2.29 |
| 2026 | 3.16 | 2.99-3.35 |
| Newborn sex | | |
| Male | REF | |
| Female | 1.22 | 1.19-1.24 |
| Maternal age | | |
| 15-18 | REF | |
| 19-29 | 1.66 | 1.43-1.93 |
| 30-39 | 1.86 | 1.60-2.16 |
| 40-49 | 2.4 | 2.05-2.81 |
| Maternal insurance | | |
| Commercial | REF | |
| Medicaid | 1.19 | 1.16-1.23 |
| Other | 0.83 | 0.78-0.88 |
| Unknown | 1.13 | 1.09-1.17 |
| Maternal ethnicity | | |
| Hispanic or Latino | REF | |
| Not Hispanic or Latino | 1.53 | 1.48-1.59 |
| Unknown | 1.69 | 1.59-1.80 |
| Maternal race | | |
| White | REF | |
| American Indian or Alaskan Native | 0.86 | 0.69-1.08 |
| Asian | 0.40 | 0.38-0.42 |
| Black | 0.78 | 0.75-0.81 |
| Native Hawaiian or Other Pacific Islander | 0.32 | 0.26-0.39 |
| Other | 1.02 | 0.97-1.07 |
| Unknown | 1.12 | 1.06-1.17 |
| Maternal residence | | |
| Urban | REF | |
| Rural | 1.02 | 0.98-1.06 |
| Unknown | 1.12 | 1.08-1.16 |

**Supplementary Table S3.** Interrupted time series model results for *receipt* of vitamin K at birth

| Variable | Adjusted Odds Ratio | 95% CI |
|---|---|---|
| Time variables | | |
| Time (months) - pre-2026 | 0.99 | 0.99-0.99 |
| January 2026 Vaccine Schedule change | 1.04 | 0.96-1.12 |
| Time (months) - post January 2026 | 0.90 | 0.88-0.91 |
| Maternal age | | |
| 15-18 | - | - |
| 19-29 | 0.60 | 0.52-0.70 |
| 30-39 | 0.54 | 0.46-0.63 |
| 40-49 | 0.42 | 0.36-0.49 |
| Maternal race | | |
| White | - | - |
| American Indian or Alaska Native | 1.15 | 0.92-1.45 |
| Asian | 2.51 | 2.36-2.66 |
| Black | 1.28 | 1.23-1.34 |
| Native Hawaiian or Other Pacific Islander | 3.10 | 2.55-3.78 |
| Other | 0.98 | 0.93-1.03 |
| Unknown | 0.90 | 0.85-0.94 |
| Maternal ethnicity | | |
| Hispanic or Latino | - | - |
| Not Hispanic or Latino | 0.65 | 0.63-0.67 |
| Unknown | 0.59 | 0.55-0.63 |
| Maternal residence | | |
| Urban | - | - |
| Rural | 0.98 | 0.94-1.02 |
| Unknown | 0.89 | 0.86-0.93 |
| Maternal insurance | | |
| Commercial | - | - |
| Medicaid | 0.84 | 0.81-0.86 |
| Other | 1.21 | 1.14-1.28 |
| Unknown | 0.88 | 0.85-0.91 |
| Newborn sex | | |
| Male | - | - |
| Female | 0.82 | 0.8-0.84 |

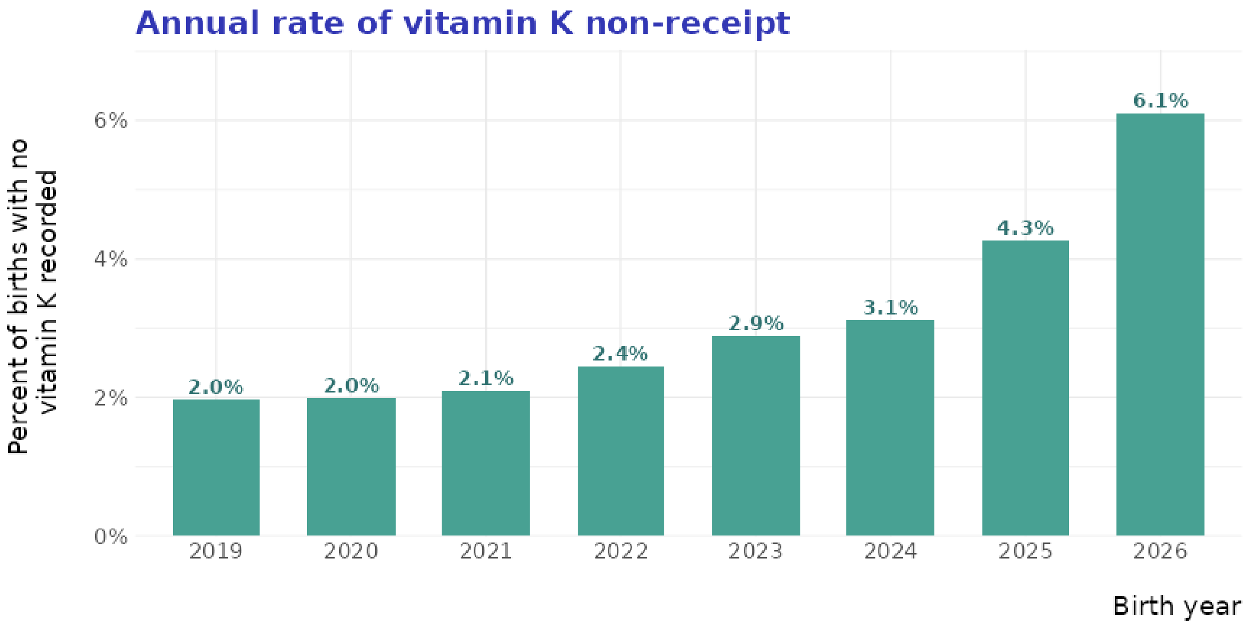

**Supplementary Figure S1**. Annual rate of vitamin K non-receipt

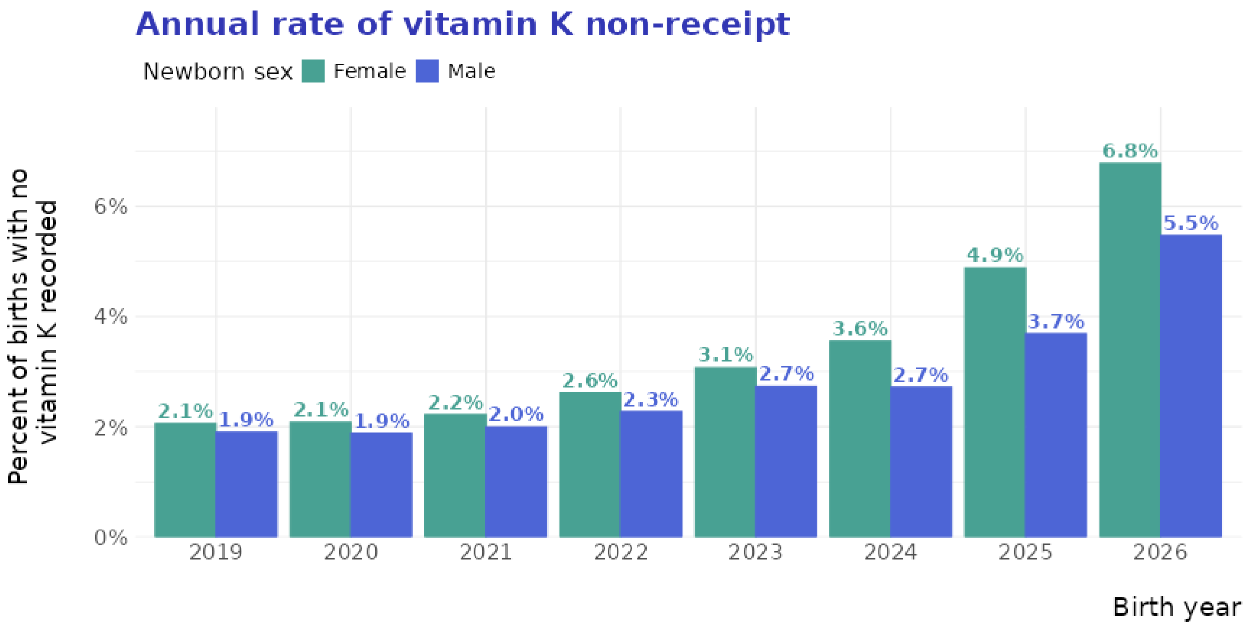

**Supplementary Figure S2.** Annual rate of vitamin K non-receipt by newborn sex

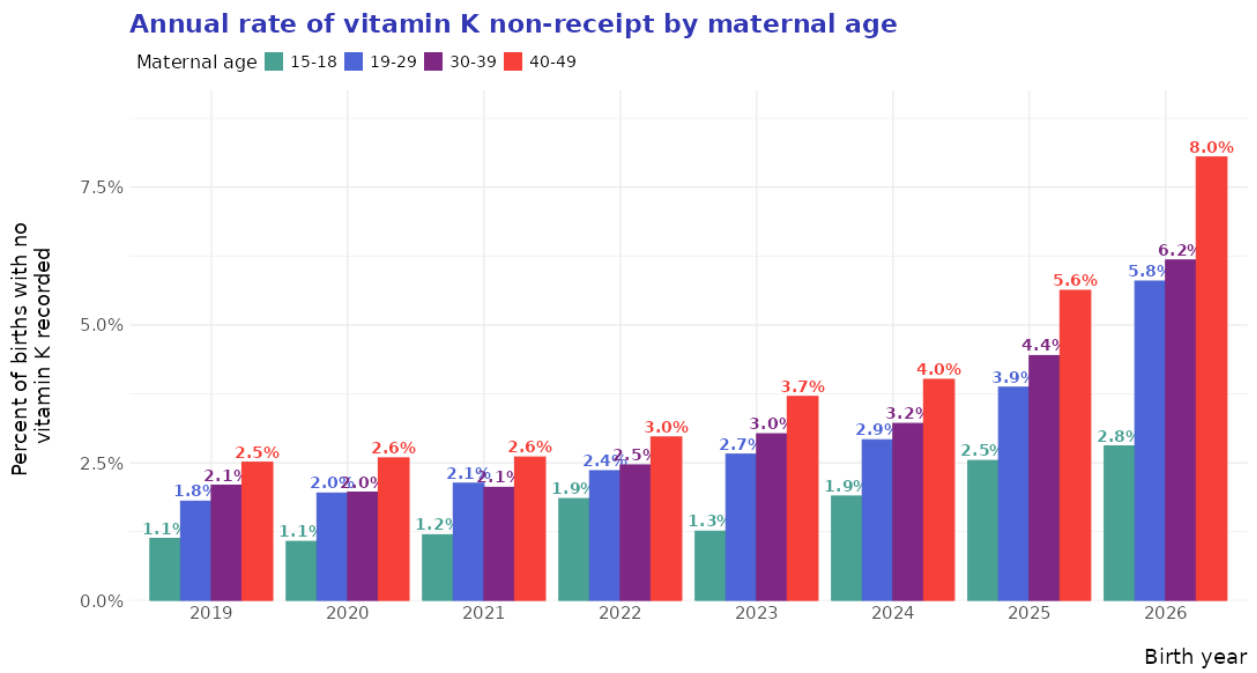

**Supplementary Figure S3.** Annual rate of vitamin K non-receipt by maternal age

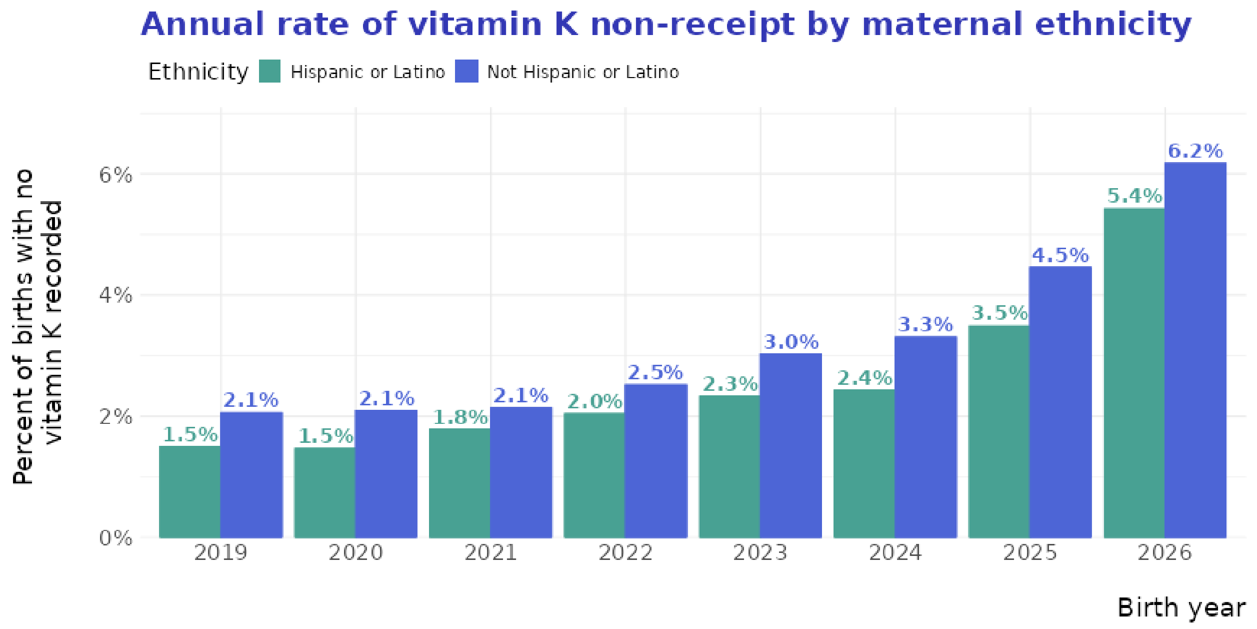

**Supplementary Figure S4**. Annual rate of vitamin K non-receipt by maternal ethnicity

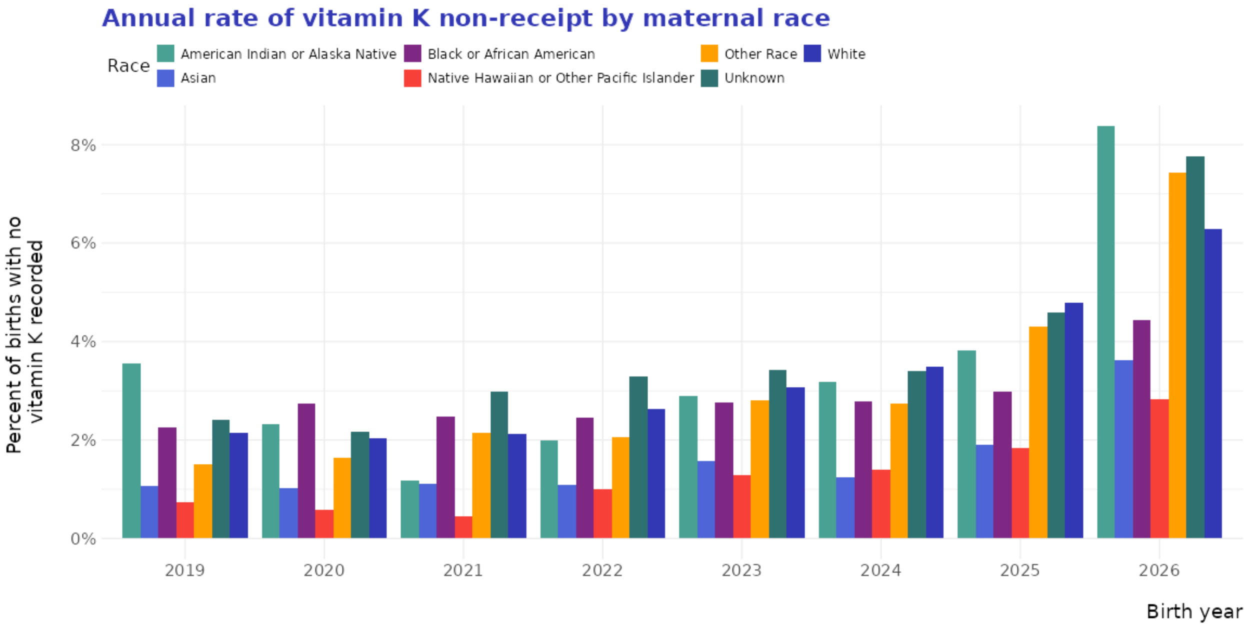

**Supplementary Figure S5**. Annual rate of vitamin K non-receipt by maternal race

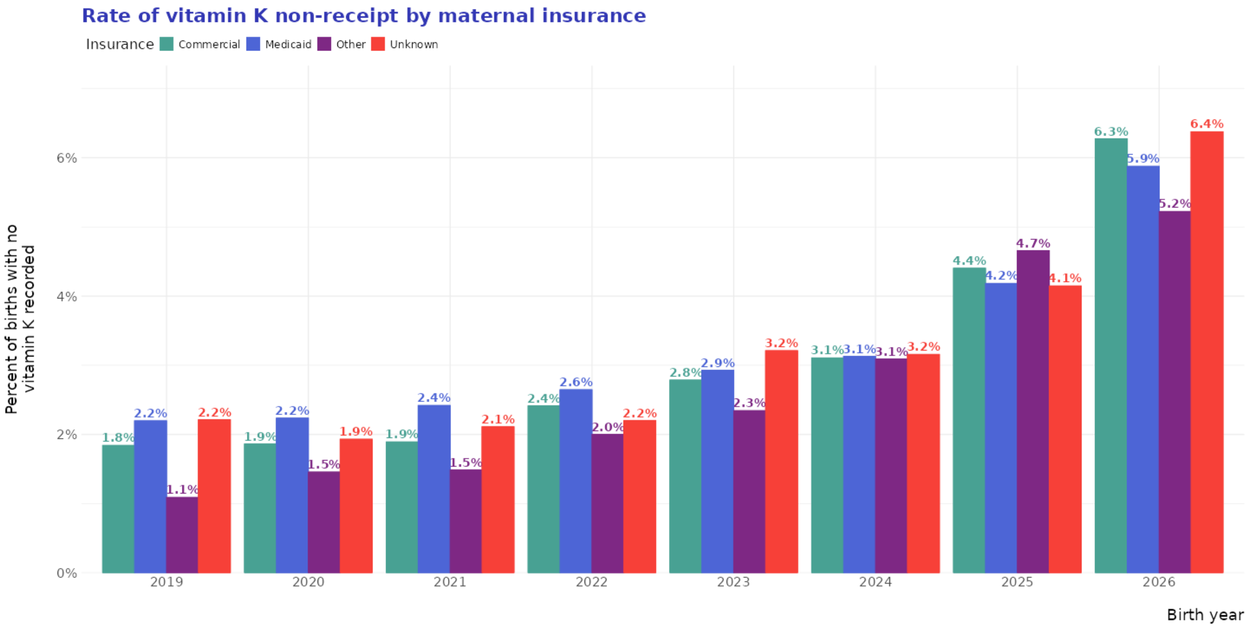

**Supplementary Figure S5.** Annual rate of vitamin K non-receipt by maternal insurance
